# Harnessing Greater Statistical Power: Comprehensive Evaluation of Disease Modifying Treatment Effects Across All or Multiple Post-Baseline Visits Compared to the Last Visit for Alzheimer’s Disease Clinical Trials

**DOI:** 10.1101/2025.02.18.25322498

**Authors:** Guoqiao Wang, Tianle Chen, John O’Gorman, Yan Li, Caiyan Li, Leonard Guizzetti, Brian Mangal, Whedy Wang, Shuang Wu, Dave Inman, Eric McDade, Randall J. Bateman

**Affiliations:** Department of Neurology, Washington University in St. Louis, St Louis, 63130, USA; Division of Biostatistics, Washington University in St. Louis, 63130, USA; Biogen Inc., 225 Binney Street, Cambridge, 02142, USA; Alector, Inc., South San Francisco, 94080, United States; Solara Consulting Corp., 2311 Swinburne Ave. North Vancouver, BC V7H 3A6, Canada; Tenaya Therapeutics, South San Francisco, 94080, United States; GlaxoSmithKline Medicines Research Centre, Stevenage, SG1 2NY, UK

**Keywords:** Alzheimer’s disease, disease-modifying, MMRM, pMMRM, Natural cubic spline, Proportional NCS, AUC, SAS

## Abstract

**Background:** In Alzheimer’s disease (AD) clinical trials, efficacy inference is traditionally based on the last visit (e.g., 18 months). However, recent studies suggest that disease-modifying treatment effects may emerge as early as 3 months post-baseline.

**Objective:** To explore this further, our study aimed to assess the increased statistical power achieved by incorporating all or multiple post-baseline visits to estimate treatment effect, compared to relying solely on the last visit.

**Methods:** We developed explicit formulas for the base functions of the natural cubic spline model, ensuring compatibility with standard SAS procedures. Through simulations using disease progression trajectories from ClarityAD and TRAILBLAZER-ALZ2 trials, we comprehensively evaluated various models in terms of power and type I error. Additionally, we offer SAS codes that to facilitate seamless implementation of different modeling approaches.

**Results:** Simulations based on ClarityAD and TRAILBLAZER-ALZ2 disease trajectories demonstrated that models incorporating multiple or all post-baseline visits yield greater power than those using only the last visit, while maintaining Type I error control. Furthermore, when three post-baseline visits were included, adding more visits resulted in minimal power gains.

**Conclusions:** Our findings support prioritizing statistical models that incorporate multiple or all post-baseline visits for treatment efficacy inference, as they offer greater efficiency than models relying solely on the last visit.

## 1. Introduction

In Alzheimer’s disease (AD) clinical trials, efficacy is usually evaluated by the treatment difference in change from baseline (CFB) of the clinical endpoints between the active treatment group and placebo group at the last study visit, which is usually 18, 24, or 27 months for early AD clinical trials. MMRM is a well-accepted statistical efficacy inference approach for AD clinical trials.^1^ Although the CFBs in each group are estimated at each study visit, only treatment difference at the last visit is used to judge if a study is positive or not. This is the current standard practice in the AD field, and henceforth we refer to this model as MMRM-Last-Visit. Some recent clinical trials for AD have indicated that disease-modifying treatment effects can begin to manifest as early as 3 to 6 months, rather than the previously considered 18 months^2, 3^. Given these findings, it is reasonable to consider models that estimate an overall treatment effect across all post-baseline visits, or across multiple prespecified post-baseline visits during the treatment period. The hypothesis posits that utilizing data from multiple study visits, rather than only the final visit, for efficacy inference can increase the efficiency of statistical tests (i.e. greater power) and, consequently, reduce the required sample size to achieve the same statistical power or increase the precision of the subgroup analyses. An additional benefit of a smaller sample size is that, when the investigational drug has worse outcomes than placebo,^4^ fewer participants are exposed to a potentially harmful drug, thereby upholding the “do no harm” principle.^5^ Furthermore, models that incorporate multiple or all post-baseline visits for efficacy inference address a more clinically meaningful question, as they evaluate the treatment effect for participants from the first included post-baseline visit through the full trial duration. In contrast, models focusing solely on the last visit assess the treatment effect only for participants who have completed the entire trial and been exposed to the treatment for its full duration. Generally, two types of models have been proposed for longitudinal data analysis to estimate treatment effects based on multiple study visits. The first type treats time as a categorical variable, using a proportional treatment effect concept such as the proportional MMRM (pMMRM) or using the adjusted mean comparison across all or multiple post-baseline visits based MMRM.^6-12^ The second type treats time as a continuous variable and includes techniques like the linear mixed effects model with a first-order continuous time.^7, 13^ Furthermore, recent studies have proposed application of natural cubic spline models as an alternative approach to evaluate the treatment effect at the final visit.^3, 14, 15^ However, it is worth noting that the publications utilizing these models did not provide clear descriptions of the natural cubic spline base functions, which can pose challenges in reproducing the analysis results.

Besides the variation in the amount of data used for the efficacy point estimates, another major difference lies in how the data being analyzed regardless of time being continuous or categorical. The common approach is to model the change from baseline with baseline being a covariate (i.e., on the right side of the regression model). The less frequently used approach is to model the post-baseline, actual values directly with baseline being part of the outcome (i.e., on the left side of the regression model). When modelling the actual value directly, the model can be set up as constrained or unconstrained. The constrained model requires that the baseline mean for the treatment and placebo group to be the same due to randomization. These variations in how to model longitudinal data warrant a comprehensive evaluation, especially in the context of positive clinical trials such as the lecanemab Clarity AD trial^2^ and the donanemab TRAILBLAZER-ALZ2 trial^3^. These clinical trials provide a unique and unprecedented opportunity for comparing model performance when investigational drugs demonstrate disease-modifying effects. The objectives of our study are to: (i) Assess the power gain achieved by estimating an overall treatment effect across all or multiple post-baseline visits compared to estimating the treatment effect solely at the last visit. (ii) Conduct a comprehensive evaluation of different statistical models by analyzing disease progression trajectories observed in these trials. (iii) Provide explicit formulas for natural cubic spline base functions that align with those utilized in standard SAS procedures. (iv) Offer detailed SAS code to facilitate the implementation and generalization of these complex models.

In Section 2, we introduce the statistical models evaluated and describe the simulation settings that reflect a range of assumptions typical of contemporary early AD trials. In Section 3, we present the simulation results, including power and type I error and discuss the performance of the models, highlighting their advantages and limitations, and provide recommendations for model selection in future studies. We concluded this study with the discussion Section 4.

## 2. Methods

Each statistical model is briefly described below. For more detailed information, please refer to the corresponding references provided for each model.

### 2.1 Statistical Models

#### 2.1.1 MMRM Based Models

Let *y*_*ijz*_ denote the longitudinal assessments for subject *i* at time *j* for treatment group *z, i* = 1, 2,…,*n, j*=0,1,…,*m*_*i*_, and *z* = 1,2 represents the placebo group and the treatment group, respectively. The marginal MMRM can be written as:

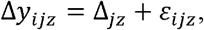

where Δ*y*_*ijz*_ is the individual change from the baseline and is calculated as *y*_*ijz*_ − *y*_*i0z*_ for *j* = 1, …, *m*_*i*_, Δ_*jz*_is the change from baseline for the group *z* at time *j* for *j* ≥ 1; *ε*_*ijz*_ is the within-subject error and is assumed to follow the same distribution for both the placebo group and the treatment group: 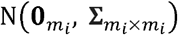, as is customarily in clinical trial analysis.^2, 3, 16^

Denote the total number of post-baseline visits as *M*, and the last study visit (e.g., 18 months) as *M* th visit. In the MMRM-Last-Visit analysis, treatment efficacy is assessed by comparing Δ_*M*l_ and Δ_*M*2._ In contrast, the MMRM-All-Visits model determines efficacy by comparing the average across all post-baseline visits, calculated as 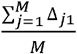 and 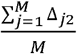. When efficacy is evaluated based on only a subset of post-baseline visits (e.g., from *m* visit to *M*) in the MMRM model, the comparison is based on the average across these specific visits, calculated as 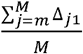 and 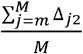. Regardless of the number of post-baseline visits used for efficacy inference, all post-baseline visits are included in the MMRM model.

Therefore, for a two-sided test, the null hypothesis of MMRM is: 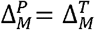 and the alternative is: 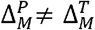. The null hypothesis for MMRM-All-Visits is: 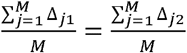, and the alternative is: 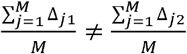.

#### 2.1.2 Proportional MMRM

The details of pMMRM and its implementation have been previously reported.^7, 12^ Briefly, it can be described as:

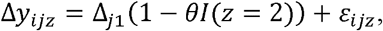

where, Δ*y*_*ijz*_ is the change from baseline for subject *i* from treatment group *z* at time *j* for *j* = 1, …., *m*_*i*_, and *z* = 1 represents the placebo group and *z* = 2 the treatment group, Δ_*j*l_ is the mean change from baseline for the placebo group at time *j* for *j* ≥ 1; *l* is the indicator function with *l*(*z* = 2) = 1 and *l*(*z* = 1) = 0; *θ* is the proportional reduction in disease progression associated with the treatment relative to the disease progression observed in the placebo group; *ε*_*ijz*_ is the within-subject error term and is assumed to follow a multivariate normal distribution: 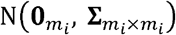.

For a two-sided test, the null hypothesis of pMMRM is: **θ** = 0 and the alternative is **θ** ≠ 0 with **θ** > 0 favoring the treatment and **θ** ≤ 0favoring the placebo.

#### 2.1.3 Linear Mixed Effects Model with First-order Time

The linear mixed effects model (LME) with first-order time as a continuous variable^17^ (i.e., random coefficient model^13^) can be described by the following equation:

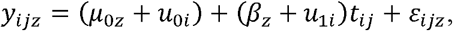

where, *y*_*ijz*_ denote the longitudinal assessments for subject *i* at time *j* for group *z, i* = 1,2,…, *n, j =* 0,1,…, *m*_*i*_, with representing the baseline visit, and *z* = 1,2 representing the placebo group and the treatment group; *μ*_0*z*_ represents the baseline mean for group *z, β*_*z*_ is the slope (e.g. the mean annual change of group *z* when time *t*_*ij*_ represents year); *β*_*z*_ *t*_*ij*_ is the mean change from baseline at time *t*_*ij*_ for group *z* assuming *t*_*i*0_ = 0, *β*_1_ − *β*_2_ represents the treatment effect; *u*_0*i*_,*u*_1*i*_ are the random effects for the intercept and the slope and are assumed to follow the same bivariate normal distribution for both groups: 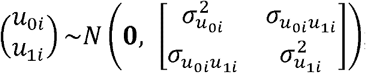 the within-subject error is assumed to follow the same normal distribution for both groups 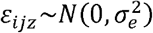. LME models have a restrictive assumption that the disease progression trajectory must be linear during the follow-up period. Consequently, this approach enforces a constant difference in the slopes (*β*_1_ − *β*_2_) at each time point and assumes a proportional treatment effect 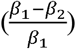 over time.

When the baseline means,, are constrained to be the same across groups ( ), reflecting the equalization effect of randomization, the LME model is commonly referred to as the constrained LME (cLME) model (Figure 1). Therefore, for a two-sided test, the null hypothesis for both LME or cLME is:, and the alternative is: .

**Figure 1:**
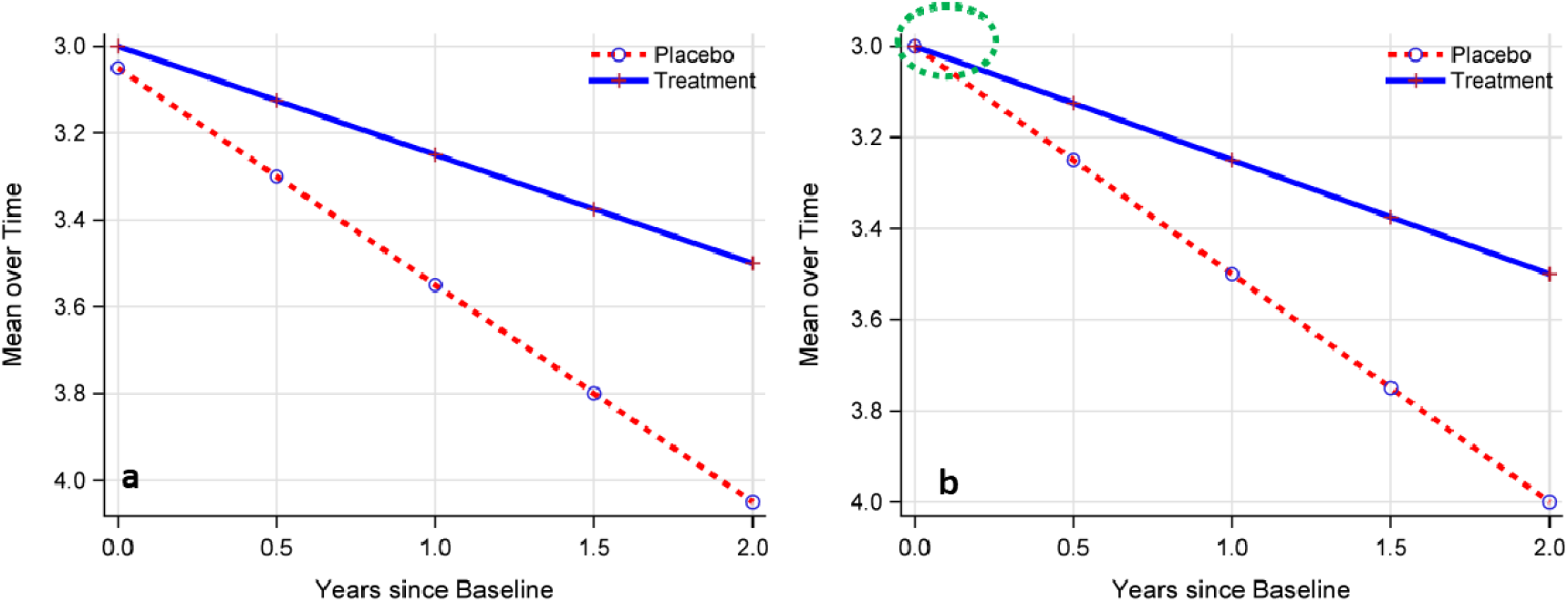
Illustration of LME (Panel a) versus constrained LME (Panel b). The green circle indicates the baseline mean is constrained to be the same for both the treatment and placebo groups.

#### 2.1.4 Natural Cubic Spline (NCS) and Constrained NCS Mixed Models

Natural cubic splines (NCS) are cubic splines that have the additional constraints that they are linear before the first knot and after the last knot.^18, 19^ Let denote the longitudinal assessments for subject at time for group, , , with representing the baseline visit, and representing the placebo group and the treatment group. For natural cubic spline of with knots , the base functions can be defined as:

Where _____, and .

In the case of 3 knots, the base function can be written as:

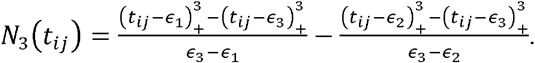

To illustrate the practical implementation of these formulas, we followed the same approach as the TRAILBLAZER-ALZ 2 donanemab trial by positioning the three knots in the time variable at the minimum, median, and maximum values.^3^ To simplify the calculation process, we divided the weeks by 4 to convert the visit schedule to months instead of weeks: 0, 3, 6, 9, 13, 16, and 19 months. Additionally, for demonstrative purposes, we assumed that all assessment times are whole numbers instead of decimal values, which may be more akin to a real trial scenario.

Similarly, following the primary analysis model setup in the donanemab trial,^3^ we positioned three knots at time points 0 (minimum), 9 (median), and 19 (maximum). As a result:

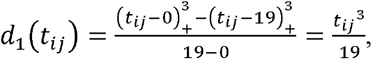

and

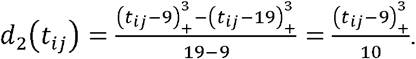

Specifically, for

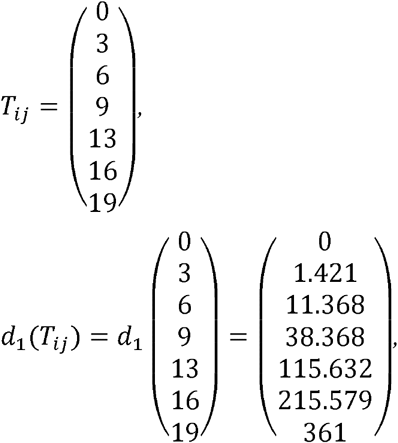

and

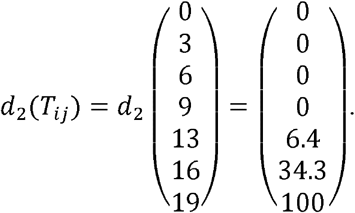

Therefore, the three NCS base functions at each time point *t*_*ij*_ can be realized as: 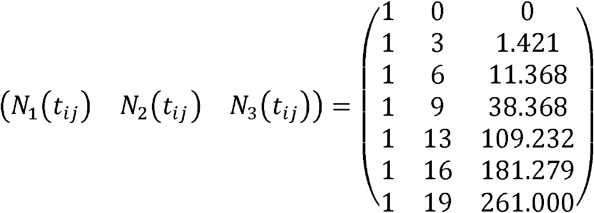, which matches the functions employed in SAS procedures such as glimmix and glmselect. In this case of positioning the knots as done in the TRAILBLAZER-ALZ 2 trial, there is essentially no restriction on fitting a linear line before the first knot (i.e., the minimum) or after the last knot (i.e., the maximum). As a result, the TRAILBLAZER-ALZ 2 trial utilized a cubic spline model rather than a natural cubic spline model.

Under a NCS model, *y*_*ijz*_ can be expressed as:

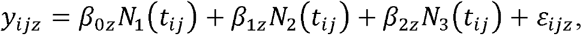

where, **β**_0*z*_ is the baseline mean for group *z, *β**_1*z*_ and **β**_2*z*_ are the coefficients associated with the linear and cubic functions of *t*_*ij*_ for group *z.ε*_*ijk*_ is the within-subject error and is assumed to follow the same distribution for both the placebo group and the treatment group: 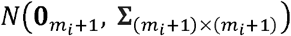, as is customarily in clinical trial analysis^2, 3, 16^.

Alternatively, if the three knots in the visit schedule of the TRAILBLAZER-ALZ 2 trial are at the 10^th^, 50^th^, and 90^th^ percentiles, which correspond to 0, 6, and 16 months after accounting for the missing data, then the same calculation process yields the three natural cubic spline functions as follows: 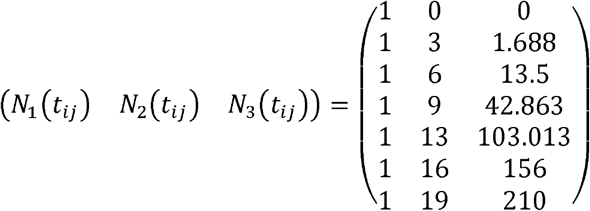, which again matches the outputs from those two SAS procedures (glimmix and glmselect).

When the baseline means, **β**_0*z*_, are constrained to be the same across groups (**β**_0_), reflecting the equalization effect of randomization, the NCS model is commonly referred to as the constrained NCS (cNCS) model.

Because NCS models the raw score directly instead of the change from baseline, there are multiple ways to express the null hypothesis. When the statistical inference is based on the mean comparison at the last visit, the null hypothesis for NCS can be written as:

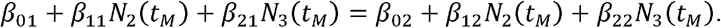

When the statistical inference is based on the change from baseline to the last visit, the null hypothesis for NCS can be written as:

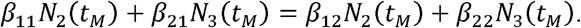

In contrast, for cNCS, the null hypothesis can be described in the same way regardless of the basis of statistical inference, as the shared baseline is cancelled out:

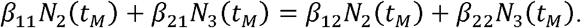

Here, *t*_*M*_ is the pre-specified time point for efficacy inference.

#### 2.1.5 Proportional NCS and Constrained Proportional NCS Mixed Models

Applying the concept of proportional treatment effect to NCS or cNCS, we obtained the following proportional NCS (pNCS)^20^:

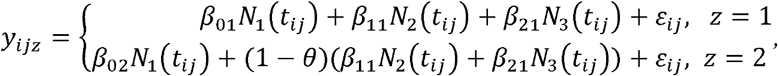

or the proportional cNCS:

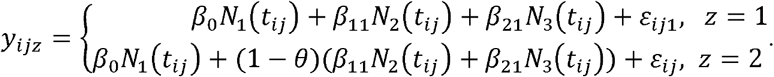

Similar to the pMMRM, pNCS estimates the treatment effect across all or multiple post-baseline visits and the null hypothesis is: **θ** = 0 and the alternative is **θ** ≠ 0 with **θ**> 0 favoring the treatment and **θ** ≤ 0 favoring the placebo.

### 2.2 Disease Progression Trajectories to Be Evaluated

As demonstrated in clinical trials with various enrollment criteria,^2, 3, 16, 21, 22^ the placebo group demonstrated relatively linear and stable disease progression trajectories over time in clinical endpoints such as CDR-SB^®^, ADAS-Cog13, ADCS-iADL, iADRS, MMSE and so on. The treatment group, however, demonstrated various trajectories depending on multiple factors such as the type of treatment effect (disease-modifying vs symptomatic), potential practice effect (i.e., learning effect),^23, 24^ or potential negative treatment effect.^21, 25^ For instance, both lecanemab and donanemab demonstrated a relatively proportional treatment effect from as early as 3 months/12 weeks, on the CDR-SB.^2, 3^ However, iADRS and its component ADAS-Cog13 exhibited temporary improvement from baseline followed by the resumption of disease progression in participants treated with donanemab.^3, 8^ Contrarily, potentially negative treatment effects can lead to faster disease progression in the treatment group compared to the placebo group.^21, 25^ Consequently, our objective is to investigate the disease progression trajectories observed in the primary endpoints of two successful clinical trials: CDR-SB in the lecanemab Clarity AD trial, and CDR-SB and iADRS in the donanemab TRAILBLAZER-ALZ 2 trial.

Specifically, three disease progression trajectories are investigated: (i) Relatively Linear Trajectory: This represents a proportional treatment effect and is demonstrated by the endpoint CDR-SB in the lecanemab Clarity AD trial. (ii) Non-Linear Trajectory with “Bumps” in the Middle: This shows large variations in the proportional treatment effect, as detailed in Supplemental Table 1. This trajectory is demonstrated by the endpoint CDR-SB in the TRAILBLAZER-ALZ 2 trial. (iii) Curved Trajectory in Earlier Post-Baseline Visits: This represents a non-proportional treatment effect, as illustrated by the endpoint iADRS in the low/medium population of the donanemab TRAILBLAZER-ALZ 2 trial.

### 2.3 Simulation Settings

As previously reported,^12^ the mean change from baseline in CDR-SB was derived from the figure of the lecanemab Clarity AD trial, except at 18 months where it was explicitly provided.^2^ The mean change from baseline in CDR-SB and iADRS was derived from the figure of the donanemab TRAILBLAZER-ALZ 2 trial (low/medium tau population), except at 76 weeks where it was explicitly provided.^3^ The covariance matrix (6 by 6) for changes from baseline was extrapolated from the estimated variance at the 18-month visit for lecanemab and at the 76-week visit for donanemab, and it is assumed to be the same for both the placebo and treatment groups. The covariance component was computed using the variances at each time point and a first-order heterogeneous autoregressive correlation matrix of 0.8. The dropout rates for each post-baseline visit in each arm were aligned with those observed in each trial.^2, 3^ The mean change from baseline and the variance-covariance matrices are provided in Supplemental Tables 2-4.

It is important to note that while these simulations provide useful datasets for achieving the objectives of our study, the results derived from them should not be directly compared to real trial data or used to compare treatments with one another. The simulated data may not fully capture the complexities and nuances of actual trial data and may lead to estimates that differ from those derived from real trial data, making direct comparisons inappropriate.^26^

The following assumptions were made for the simulations:

- Baseline values: Simulated using the mean (SD) published in each trial’s demographic table, constrained by the range of each endpoint.
- Post-baseline individual level data: Simulated using the mean and related variance-covariance matrix derived from published figures. Therefore, the mean treatment effect is identical to those observed in each real trial at each post-baseline visit. When evaluating type I error, both the treatment group and the placebo group were simulated based on each trial’s placebo mean decline trajectory.
- Sample size: Two sample sizes are presented for each endpoint to provide variation in power estimation while maintaining conciseness.
- Duration and visit schedule: Identical to each real trial.
- Dropout rate: Identical to each real trial at each post-baseline visit.
- Analysis strategy: All models described in Table 1 were applied to evaluate model performance for each simulated dataset. for each model a two-sided test was used for hypothesis testing, with a type I error rate set at 0.05.
- 2000 replicates were simulated for each scenario.
- Power or type I error: Calculated as the proportion of the 2000 simulated trials with a p-value less than 0.05.
- All simulations and analyses were performed using SAS procedures.

**Table 1:**
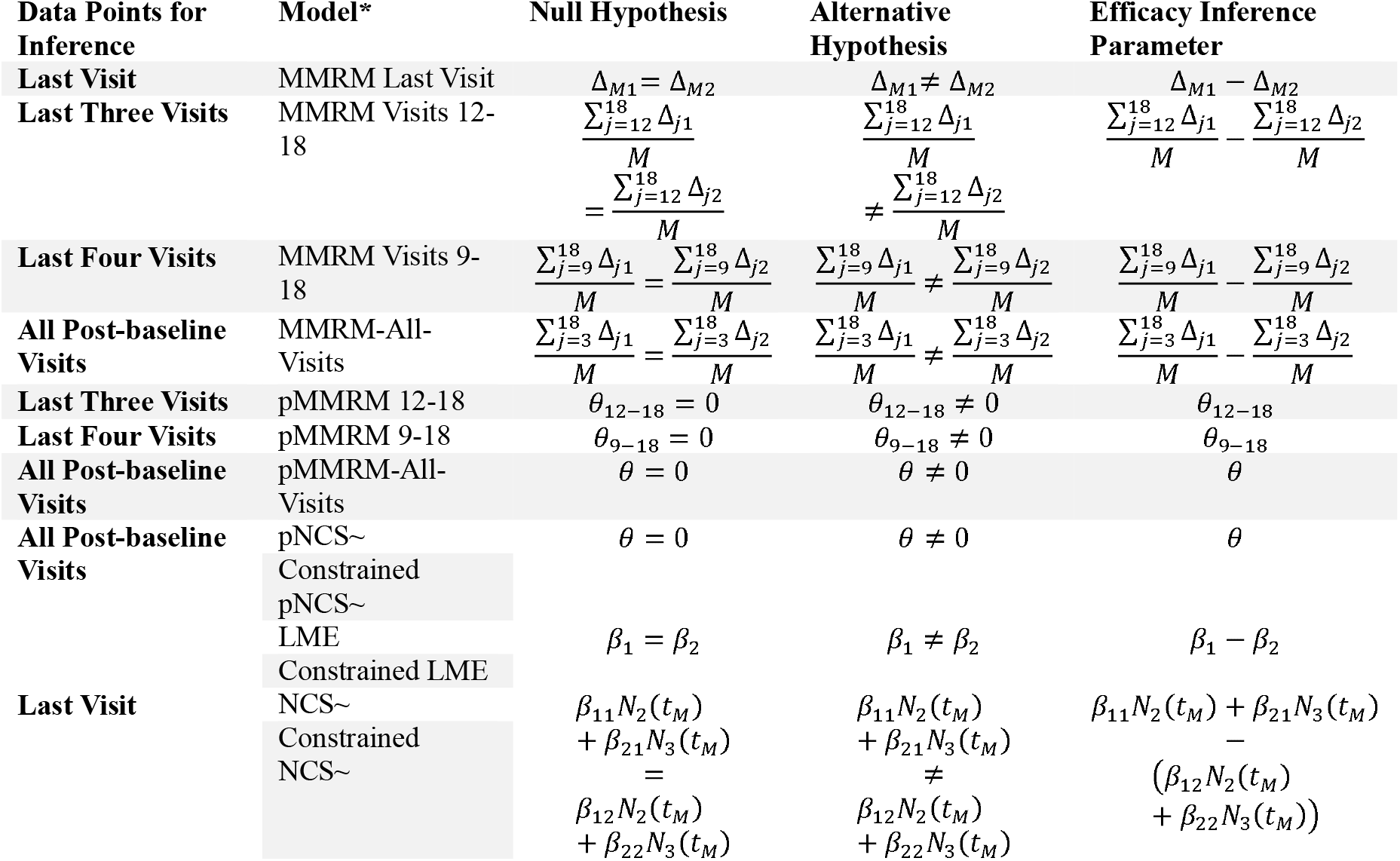
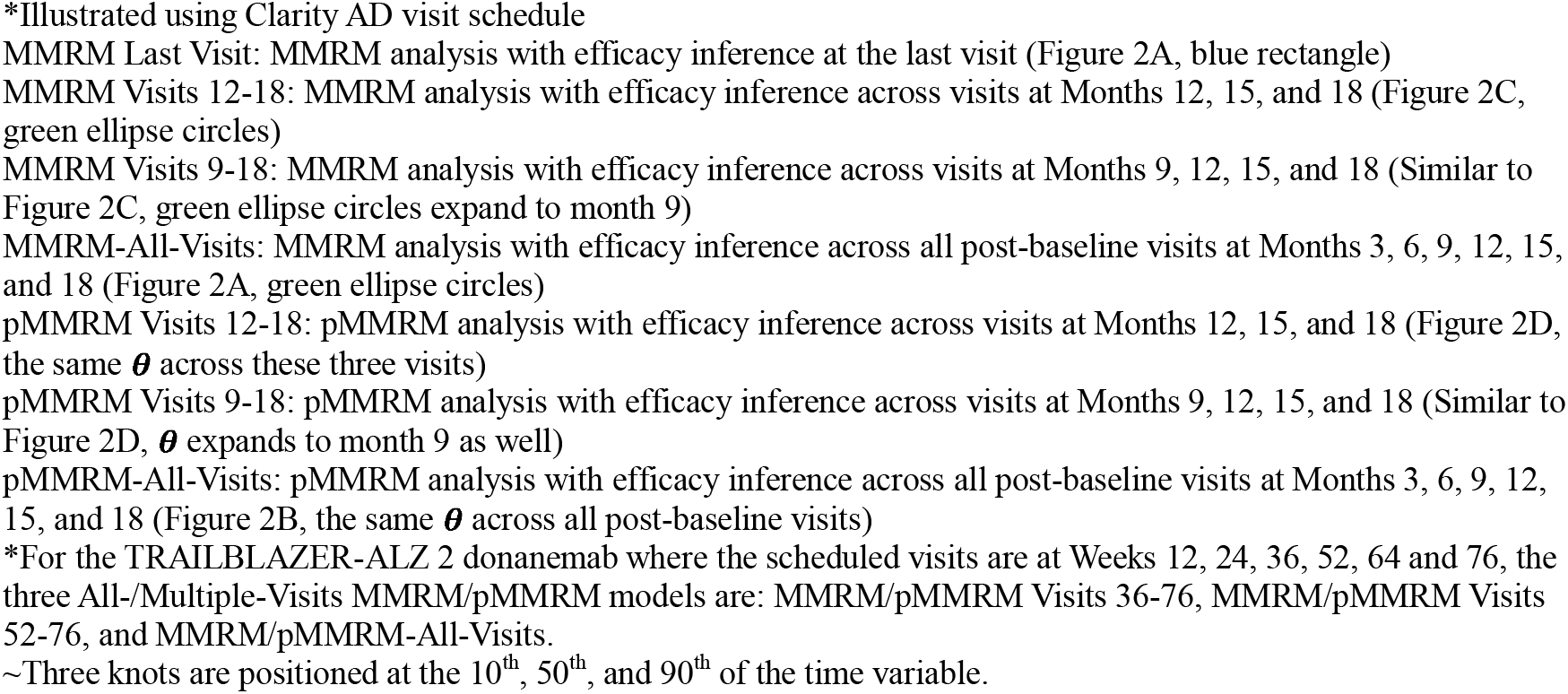
Summary of Evaluated Statistical Models and Statistical Representation of Testing Hypotheses.

It is worth noting that depending on the data points used in the efficacy inference, these models can be divided into two categories regarding the hypothesis being tested. For models that focus on the last visit, the hypothesis testing evaluates the treatment effect for participants who have been exposed to the treatment for the full duration of the trial (e.g., 18 months). For models that employes multiple or all post-baseline visits, the hypothesis testing evaluates the treatment effect for participants from the first included post-baseline visit up to the full duration of the trial. For example, if the 12-month visit is the first post-baseline visit included in the efficacy inference, the hypothesis would cover the exposure duration from 12 months to 18 months.

## 3. Results

### 3.1 Illustration of Statistical Inference Using All or Multiple Visits

Figure 2 depicts the distinctions between single-endpoint statistical inference and All- or Multiple-Visits statistical inference, as well as similar representations of pMMRM reported in previous studies.^12^ The traditional approach to efficacy inference at the last study visit, denoted by the blue rectangle, even though all post-baseline data are incorporated in the model. Alternatively, one can utilize all post-baseline visits (shown in Figure 2a and 2b) or multiple post-baseline visits (shown in Figure 2c and 2d). The dashed circle visits in Figure 2a represent the MMRM-All-Visits analysis, while the ones in Figure 2c represent the MMRM-Multiple-Visits analysis specifically conducted at Months 12, 15, and 18. The shared parameter **θ** across all post-baseline visits is demonstrated in Figure 2b, representing the pMMRM across all post-baseline visits. On the other hand, the shared parameter across months 12 to 18 represents the pMMRM within those three months (Figure 2d). Although only two scenarios (All visits vs. three visits) are illustrated in Figure 2, any number of post-baseline visits can be used for efficacy inference and illustrated in a similar manner since all post-baseline visits are included in the model.

**Figure 2:**
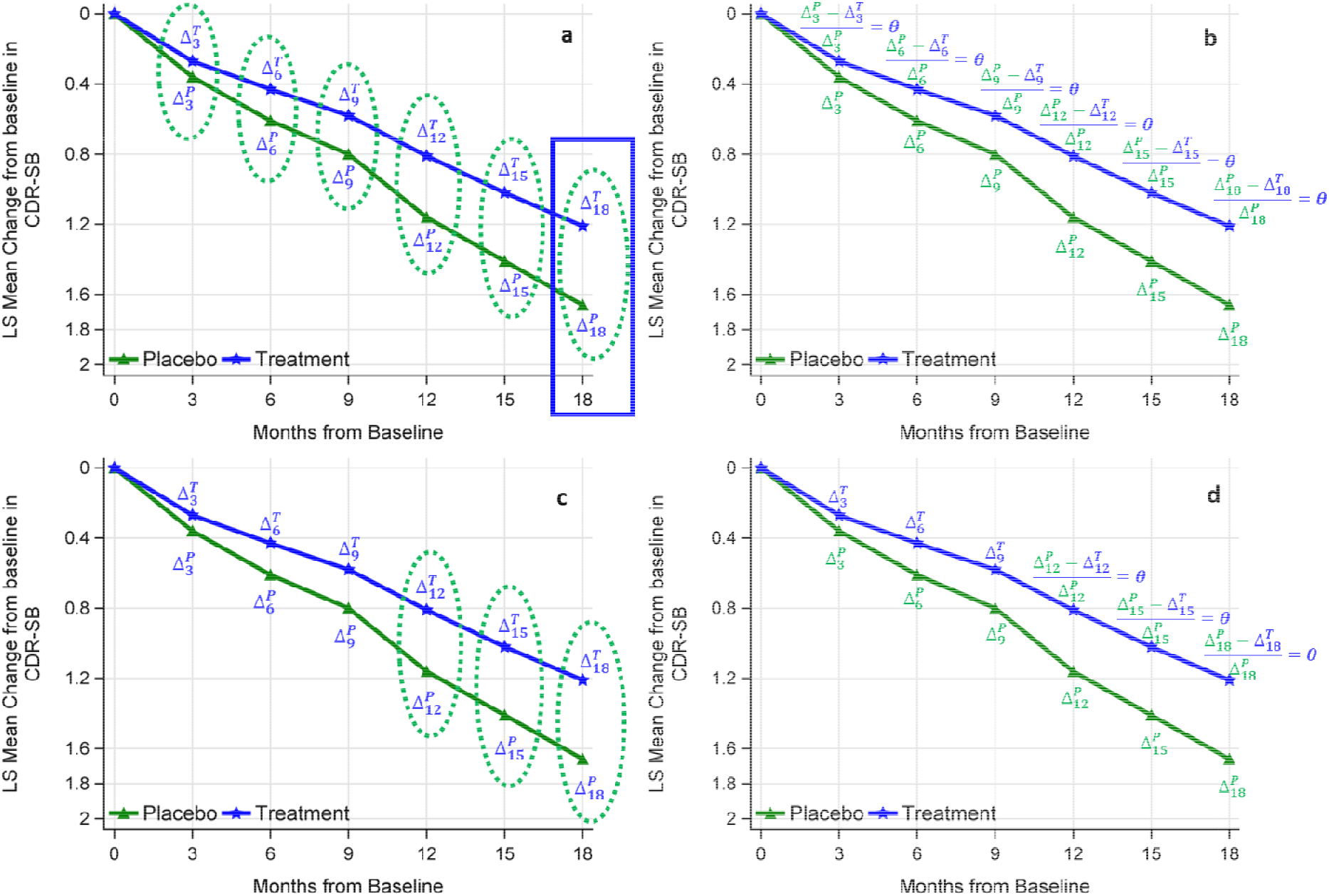
Illustration of statistical inference using different numbers of post-baseline visits. Panel a: MMRM-Last-Visit with treatment effect estimated at the last visit (blue rectangle), and MMRM-All-Visits with treatment effects estimated across all visits (green circles). Panel b: pMMRM-All-Visits with treatment effect estimated across all visits. Panel c: MMRM-Multiple-Visits with treatment effect estimated across last 3 visits (green circles). Panel d: pMMRM-multiple-Visits with treatment effect estimated across last 3 visits.

It is important to note that when pMMRM assumes the same proportional treatment effects across all or multiple visits, it does not imply that the observed proportional treatment effect (i.e., % reduction relative to placebo decline) at each post-baseline visit needs to be identical. Supplemental Table 1 presents the estimated proportional treatment effects at each visit and a comparison of those with substantial differences. Although there are numerical differences in proportional reductions across visits in the observed data, these differences are not statistically significant. This example demonstrates that the proportionality assumption is robust to the presence of variation in the observed proportional treatment effects at each visit.

### 3.2 Simulation Results based on the semi-real setting of Lecanemab Clarity AD Trial

Figure 3 illustrates the comparison of power and type I error among different models based on the disease progression trajectories of CDR-SB observed in the lecanemab Clarity AD trial. These trajectories demonstrate a relatively linear progression of the disease.

**Figure 3:**
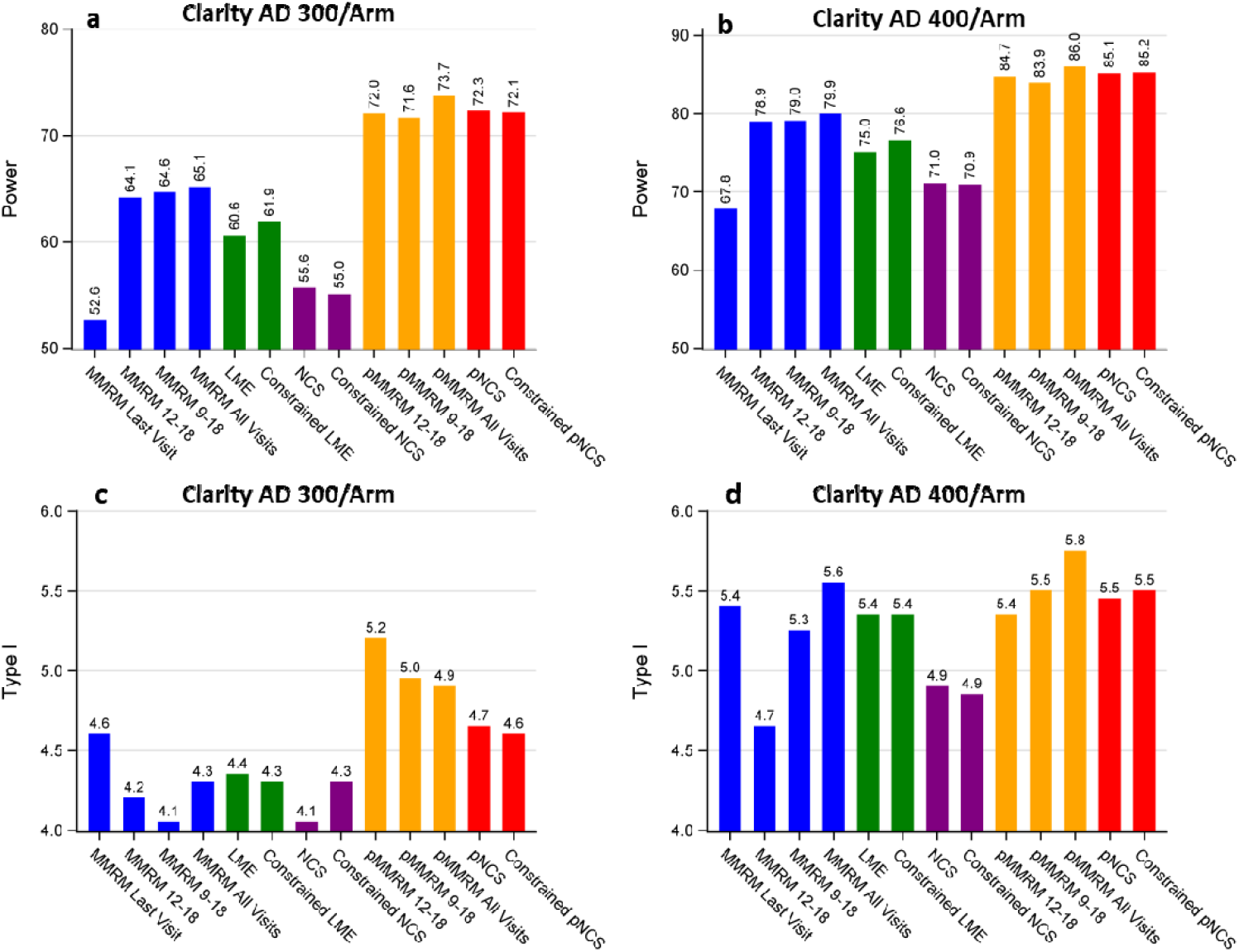
Power and Type I error comparison for different models across sample sizes based on CDR-SB. Panel A: Power comparison at a sample size of 300 per arm. Panel B: Power comparison at a sample size of 400 per arm. Panel C: Type I error comparison at a sample size of 300 per arm. Panel D: Type I error comparison at a sample size of 400 per arm.

Based on Figure 3, several observations can be made regarding power (panels a and b): (i) All other models demonstrate a power advantage over the traditional MMRM-Last-Visit analysis. (ii) Models that utilize all post-baseline visits (pMMRM-All-Visits, MMRM-All-Visits, pNCS, constrained pNCS, LME, constrained LME) or multiple post-baseline visits (pMMRM 12-18, pMMRM 9-18, MMRM 12-18, MMRM 9-18) for estimating treatment effect exhibit higher power compared to those based solely on the last visit (MMRM-Last-Visit, NCS, and constrained NCS), with proportional models showing much higher power. (iii) The use of constrained baseline results in limited power discrepancy when both the constrained and non-constrained models test efficacy based on the same metric, such as slope in LME and change from baseline in NCS. (iv) The additional post-baseline visits have a limited impact on power when the last three visits have been included in the MMRM or pMMRM model. (v) Despite differences in the number of model parameters to estimate, pMMRM and pNCS models exhibit similar power.

Furthermore, the type I error for all models appears to be controlled and mostly fluctuates within 1% of the nominal level of 5% (panels c and d).

### 3.3 Simulation Results based on the semi-real setting of donanemab TRAILBLAZER-ALZ 2 trial

#### 3.3.1 Simulation Results based on CDR-SB

Figure 4 illustrates the power and type I error comparison among different models under investigation based on the disease progression trajectories of CDR-SB observed in the donanemab TRAILBLAZER-ALZ 2 trial (low/medium tau population). Although these trajectories also demonstrate a relatively linear progression of the disease, the percentage reduction peaked at week 36 (as shown in Supplemental Table 1) and gradually declined from week 36 to week 76.

**Figure 4:**
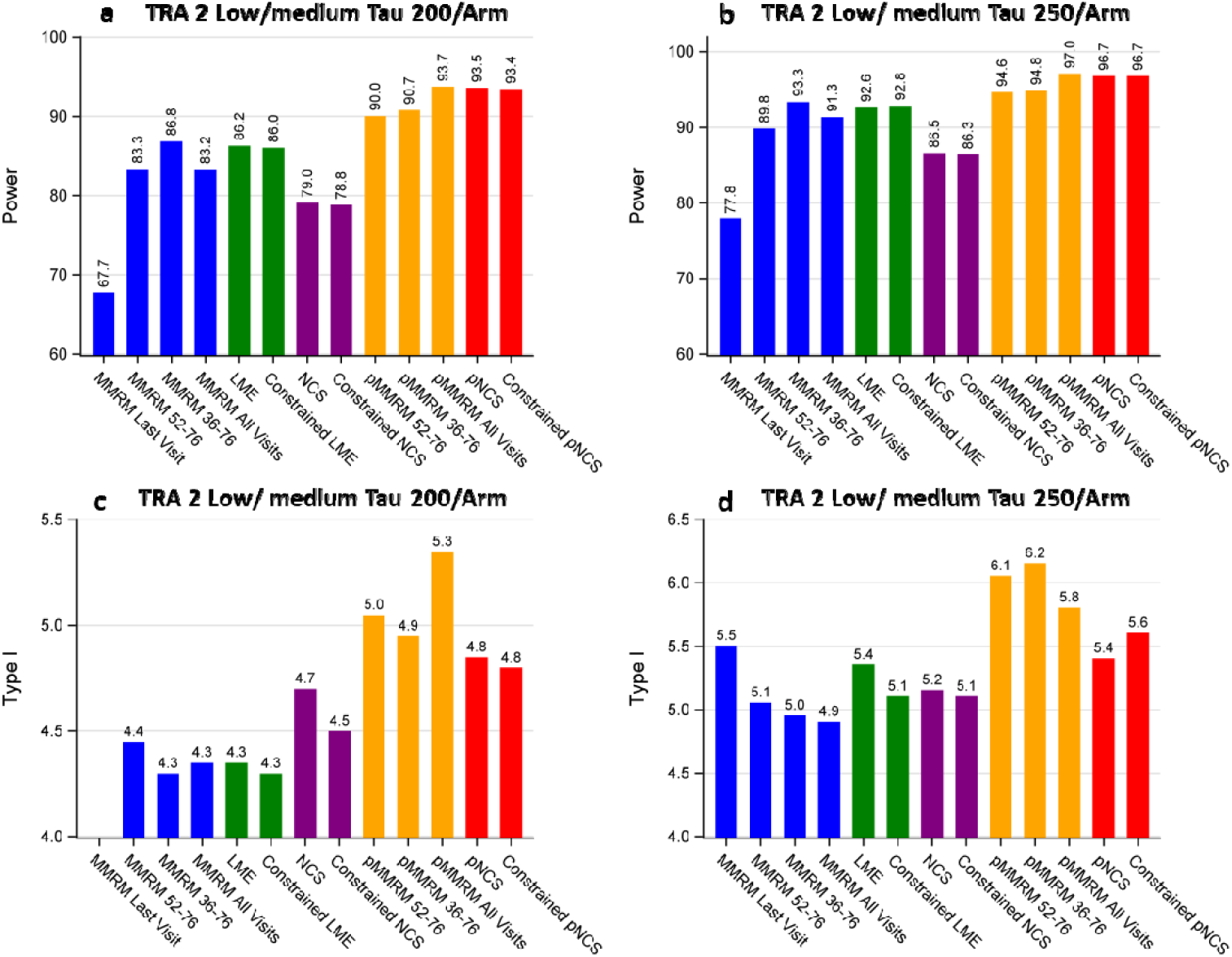
Power and Type I error comparison for different models across sample sizes based on CDR-SB. Panel A: Power comparison at a sample size of 200 per arm. Panel B: Power comparison at a sample size of 250 per arm. Panel C: Type I error comparison at a sample size of 200 per arm. Panel D: Type I error comparison at a sample size of 250 per arm. TRA 2: TRAILBLAZER-ALZ 2 Trial

Figure 4 demonstrates consistent findings with Figure 3 regarding power (Figure 4, panels a and b), while highlighting a significant difference in the MMRM related models. Notably, the separation in terms of % reduction between groups peaked at week 36, resulting in the MMRM 36-76 model displaying the highest power compared to the MMRM 52-76 or MMRM-All-Visits models. Again, the type I error for all models appears to be controlled and mostly fluctuates within 1% of the nominal level of 5% (Figure 4, panels c and d).

#### 3.3.2 Simulation Results based on iADRS

The observed disease progression trajectories of iADRS for the low/medium tau population indicate an improvement (i.e., positive change compared to baseline) in the treatment group at Weeks 12 and 24. However, starting from week 36, there is a decline in the treatment group, reflected by negative changes in the iADRS scores. In contrast, the placebo group consistently shows declines in the iADRS scores across all post-baseline visits. This improvement in the treatment group at Weeks 12 and 24 invalidates the proportionality assumption in both pNCS and pMMRM. Since the decline in the treatment group is set to be percentage of that in the placebo group, the proportional model fails to reflect the observed improvement in the treatment group. The NCS model with three knots also fails to capture this improvement in the treatment group. Figure 5 illustrates a comparison of disease progression trajectories estimated by MMRM and pMMRM, as well as a comparison by MMRM and constrained NCS. Despite violating the proportionality assumption, all three pMMRM models yield more conservative estimates in the following ways: (i) the estimated disease progression trajectories fall within those estimated by MMRM, and (ii) the estimated means of change from baseline to week 76 closely align with those estimated by MMRM. Similarly, NCS also closely aligns with MMRM at week 76.

**Figure 5:**
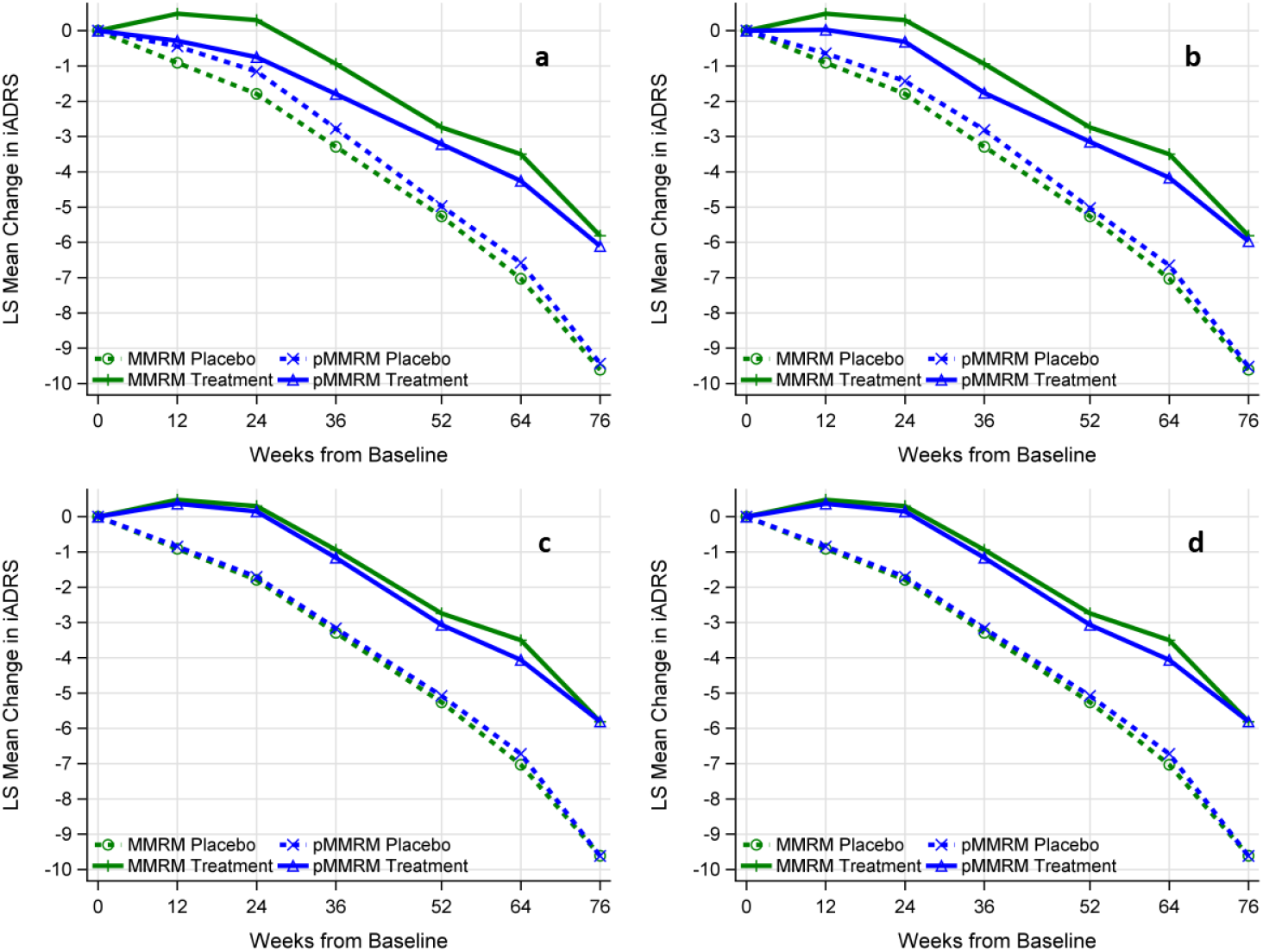
Comparison of the estimated disease progression trajectories by different models. Panel a: MMRM versus pMMRM-All-Visits with treatment effects estimated across all visits. Panel b: MMRM versus pMMRM 36-76 with treatment effect estimated across visits from week 36 to week 76. Panel c: MMRM versus pMMRM 52-76 with treatment effect estimated across visits from week 52 to week 76. Panel d: MMRM versus Constrained NCS. Panels a, b, c were generated based on a single simulated semi-real trial data, closely mimicking the real trial data. Panel d was generated by extracting the disease progression trajectories from the TRAILBLAZER-ALZ 2 trial.

Figure 6 illustrates the power and type I comparison among different models under investigation based on the disease progression trajectories of iADRS observed in the TRAILBLAZER-ALZ 2 donanemab trial (low/medium population). Several key observations can be made regarding power (Figure 6 panels a and b): (i) The violation of the proportional assumption in the pMMRM-All-Visits leads to a more conservative power, resulting in slightly lower power compared to the MMRM-Last-Visit model. The pMMRM-All-Visits model shows similar power to the NCS models. (ii) The pMMRM 52-76 model assumes the same proportional treatment effect among the last three visits and does not violate the proportionality assumption.

**Figure 6:**
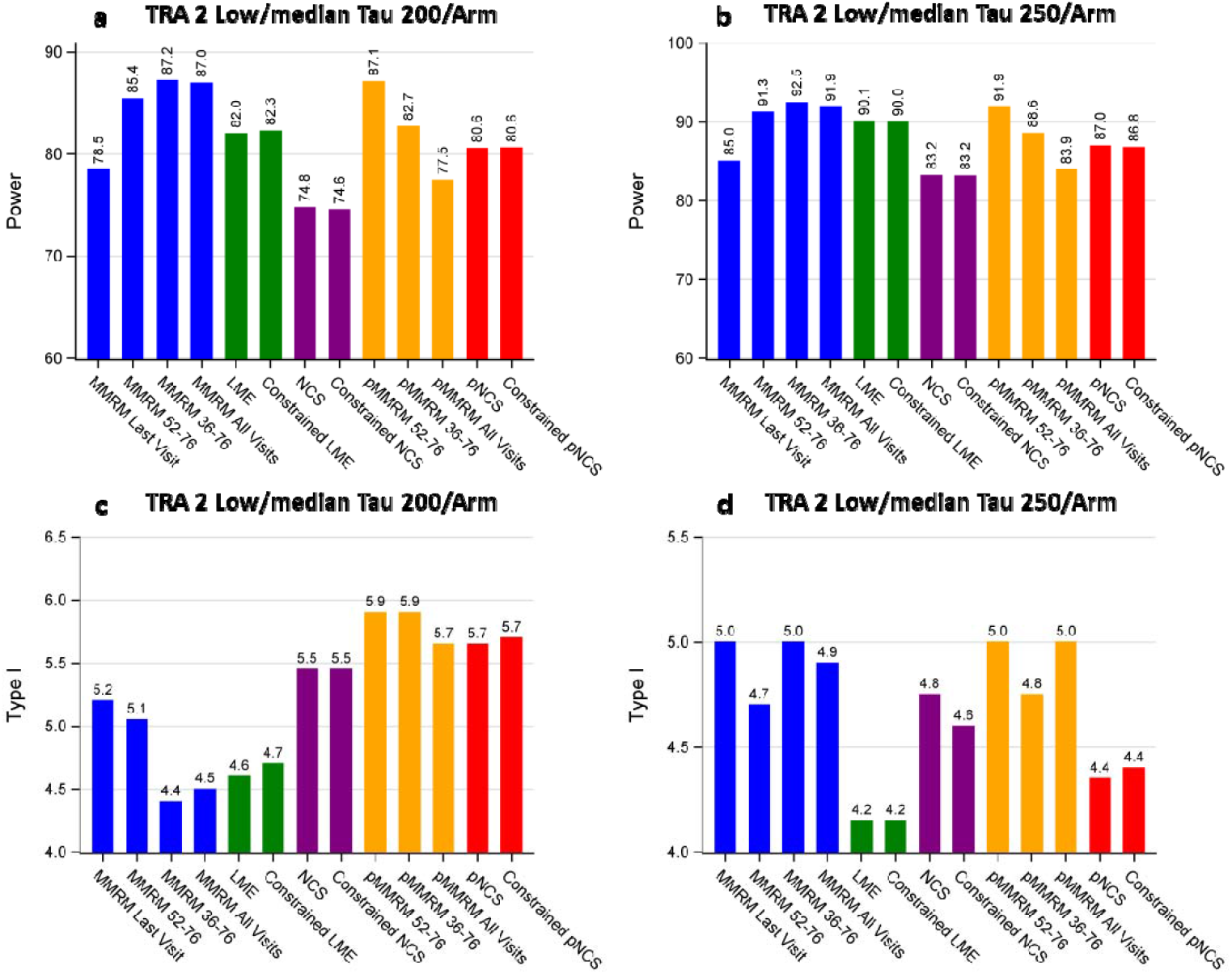
Power and Type I error comparison for different models across sample sizes based on iADRS. Panel a: Power comparison at a sample size of 200 per arm. Panel b: Power comparison at a sample size of 250 per arm. Panel c: Type I error comparison at a sample size of 200 per arm. Panel d: Type I error comparison at a sample size of 250 per arm. TRA 2: TRAILBLAZER-ALZ 2 Trial

Consequently, it exhibits a substantial increase in power compared to the MMRM-Last-Visit model. (iii) The MMRM-All-Visits or MMRM with multiple visits demonstrate the highest power by leveraging the significant separation observed in the disease progression trajectories, resulting from the improvement over baseline observed in the treatment group. (iv) Despite the non-linear disease progression trajectory in the treatment group, the constraint for the baseline mean has no impact on power for both LME and NCS models. (v) The type I error for all models appears to be controlled and mostly fluctuates within 1% of the nominal level of 5% (Figure 6 panels c and d).

### 3.4 Interpretation of Analysis Results from MMRM-All/Multiple-Visits

MMRM-All/Multiple-Visits estimates the average mean changes from baseline for both the treatment and placebo groups and makes the efficacy inference based on the difference between these mean changes. Because the difference is averaged across all or multiple visits, interpreting its clinical meaningfulness can be challenging. To facilitate interpretation, the difference can be converted to a percentage/proportional reduction relative to the average placebo mean change as:

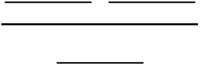. Then, it can be interpreted as the average percentage reduction of in disease progression for treated participants compared to those on placebo after up to 18 months of exposure to the treatment.

### 3.5 Compare Area Uder the Curve (AUC)

#### 3.5.1 MMRM AUC Comparison

MMRM AUC comparison evaluates whether or not the area between the disease progression trajectories is 0 and the area between the trajectories is calculated as:

from baseline to month 3: 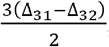,

from month 3 to month 6: 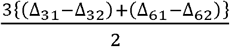,

from month 6 to month 9: 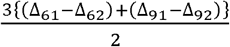,

from month 9 to month 12: 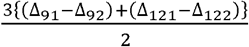,

from month 12 to month 15: 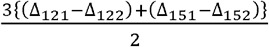,

from month 15 to month 18: 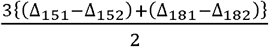.

From baseline to month 18 is the sum of all the above, after simplified it, the area between the trajectories is:

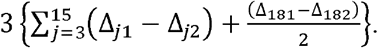

Ander the null hypothesis:

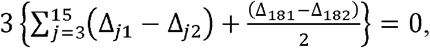

Under the alternative:

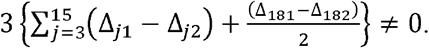

The MMRM AUC comparison produced results similar to MMRM-All-Visits and can be interpreted similarly by converting it to a percentage or proportional reduction relative to the placebo AUC (results not shown).

#### 3.5.2 NCS AUC Comparison

Another approach to comparing AUC involves using the NCS mixed model by integrating over the entire trial duration. In the example from Section 2.1.4, where knots are set at 0 (minimum), 9 (median), and 19 (maximum), AUC comparison for non-constrained NCS is performed by integrating over each group’s trial duration, with the difference calculated as follows:

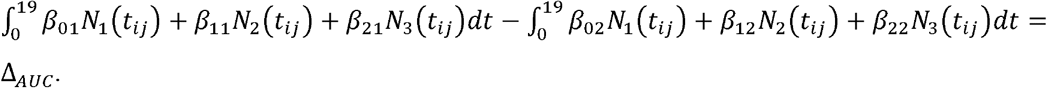

After integration, the AUC for the placebo group is:

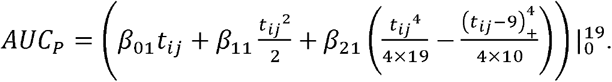

And the AUC for the treatment group is:

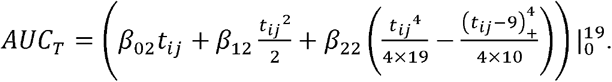

Ander the null hypothesis:

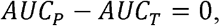

Under the alternative:

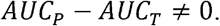

For constrained NCS, where **β**_01_ = **β**_02_, AUC comparison follows a similar approach, with the first term involving **β**_01_ and **β**_02_ canceling out.

Similarly, NCS AUC comparison can be interpreted similarly by converting it to a percentage or proportional reduction relative to the placebo AUC.

### 3.6 Recommendation of the choice of the primary analysis model

Several factors should be considered when selecting the primary analysis model for a clinical trial. First, it is important to determine whether the treatment is intended to be disease-modifying or symptomatic. Disease modifying treatments, such as anti-amyloid therapies, typically exhibit their effects early on, which could be as early as 3 months post-baseline, and these effects tend to increase over a reasonable follow-up duration, such as 18 months.^2, 3^ In contrast, symptomatic treatments tend to have diminishing effects over the 18-month period, resulting in smaller proportional treatment effects at later visits and potential violation of the proportionality assumption.

Secondly, the presence of a practice effect (i.e., learning effect) on the primary endpoint should be considered.^3, 23, 24^ If there is a notable practice effect that leads to improvement over baseline, it may invalidate the proportionality assumption in the pMMRM-All-Visits model or the linearity assumption in the LME model. Proportional models are suitable for disease modifying treatments and endpoints with minimal or no practice effects, such as CDR-SB. However, if the endpoint exhibits improvement over baseline in the earlier post-baseline visits, the MMRM-All-Visits model has the greatest power. Additionally, when practice effects exist at early visits, pMMRM with the last multiple visits (e.g., the last three visits) may have greater power compared to pMMRM-All-Visits. Therefore, pMMRM with the last multiple visits can be considered as the primary analysis model under practice effect scenarios. It is important to pre-specify in the Protocol and SAP which post-baseline visits will be included for primary efficacy inference.

Another major consideration is the goal of the trial, including the enrollment targets. For example, in a large phase 3 confirmatory trial, overall power is typically sufficient to detect the clinical effect sizes being sought. However, if the trial is designed to compare subgroups—such as APOE4 status, sex, comorbidity, or race—to determine drug effects, additional power may be necessary to increase the precision of the subgroup analyses. Similarly, in Phase II trials with relatively small sample sizes, additional power may be needed to prevent Type II errors of progression forward to phase 3 trials.

The flexibility offered by MMRM or pMMRM-based models to make efficacy inferences based on all or multiple post-baseline visits makes them highly appealing in the circumstances observed from the Clarity AD trial and the TRAILBLAZER-ALZ 2 trial. Models that leverage treatment effects across all or multiple post-baseline visits provide a substantial power advantage over those relying solely on the last study visit, and thus should be prioritized.

It is important to note that when the treatment does not have a disease-modifying effect and the disease progression trajectories either converge (donepezil),^27^ overlap (DIAN-TU, gantenerumab),^21^ or diverge unfavorably (A4, solanezumab),^25^ the proportionality assumption is inevitably violated. Thus, when the treatment is ineffective, the model assumptions become irrelevant, and the choice of models with stronger assumptions does not impact the overall statistical inference. When the treatment indeed has a disease-modifying effect, such as lecanemab or donanemab, the application of pMMRM or MMRM with all or multiple visits can significantly reduce the required sample size, therefore accelerating the development of new treatments. However, the final sample size must also consider the minimum number of subjects required to be exposed to a new investigational drug, as recommended by ICH guidelines.^28^

## 4. Discussion

The emergence of disease-modifying treatments like lecanemab and donanemab for neurodegenerative diseases such as Alzheimer’s disease presents an unprecedented opportunity to re-evaluate various statistical models. These treatments have provided insights into the disease progression trajectories under disease-modifying effects, demonstrating three unique features: (i) a continuous increase in the difference between the treatment and placebo groups in post-baseline disease progression trajectories, (ii) relatively stable proportional/percentage reduction across all or multiple post-baseline visits, and (iii) treatment effects manifesting as early as three months post-baseline. These features support the utilization of more post-baseline visits beyond the last visit for robust statistical efficacy inference. While all models listed in Table 1 incorporate all post-baseline data, it’s important to note that the data collected before the last visit do not directly contribute to the efficacy comparison in models that focus solely on the last visit for statistical inference.

Basing statistical inference on more than just the last visit offers several advantages. Firstly, estimating an average treatment effect throughout the trial duration, or a part of it, reduces the impact of survivor bias that may arise at the last visit. Secondly, it increases statistical efficiency by utilizing data from participants who dropped out before the last visit, which can account for a significant proportion of the total sample size (e.g., 16.9% in Clarity AD, 22.3% and 31.8% in the TRAILBLAZER-ALZ 2 low/medium and High-tau population, respectively). In contrast, participants who drop out early do not directly contribute to efficacy inference in models that rely the comparison solely on the last visit, such as the MMRM-Last-Visit model. Thirdly, the estimated treatment effect becomes more representative by including and utilizing all participants who were exposed to the treatment and had at least one post-baseline assessment. This approach more accurately reflects real-world observations, as it estimates the treatment effect for the intended estimand rather than relying solely on participants who received treatment throughout the entire trial duration. Overall, utilizing more than just the last visit offers several advantages including reducing survivor bias, increasing efficiency of statistical inference, and providing a more representative estimate of treatment effects. This approach better mirrors real-world scenarios where treatment durations may vary for individual patients.

In this study, we conducted a comprehensive evaluation of various models used for analyzing clinical trials with longitudinal data. We provided explicit formulas for generating the base function of NCS models. The use of these formulas enables the matching of base functions generated by standard SAS procedures, facilitating the validation and reproducibility of NCS models under any settings. Prior publications on NCS models did not provide explicit formulas, making evaluation and reproduction of these models challenging.^9, 14^

Next, we assessed the performance of these models in terms of type I error control and power. All models, including the proportional model, demonstrated control of the type I error. The simulated type I error consistently fluctuated around the nominal 5% level, which is consistent with previous reports on this topic.^7, 19^ A previous study highlighted potential issues with type I error control in a specific type of frequentist proportional model.^14^ However, several factors might have contributed to the apparent lack of control in that study. Firstly, the previous study involved directly translating a Bayesian proportional model to a frequentist model,^6^ which posed challenges regarding type I error control. The original Bayesian model utilized a pre-specified threshold of 99.52% derived from extensive simulations, instead of relying on the nominal 97.5% threshold to control one-sided type I error at 2.5%.^6^ Consequently, directly converting it to a frequentist model with the nominal 5% level for controlling two-sided type I error becomes problematic. To address this, when extending the original Bayesian proportional model to frequentist proportional models, we adjusted the model parameter setting for the proportional treatment effect to more appropriately control two-sided type I error at the nominal 5% level (from testing *e*^**θ**^ = 1 to testing **θ** = 0 ).^6, 7, 12, 26^ Secondly, the previous study encountered severe convergence issues with the computational package used, which undermined the reliability of its conclusions. When a significant percentage of models fail to converge, not only do the results from those models become unreliable, but even the results from models without convergence issues may fail to reach the global minimum point and yield accurate estimations. This could explain the scenario where the type I error was larger than the power observed in the previous study.^14^ In our simulations, we primarily utilized the SAS computational package “%nlinmix” macro, and convergence issues were not encountered. SAS procedure “nlmixed” can also be used to implement these models,^12^ but we prefer “%nlinmix” macro for its convenience in incorporating various variance-covariance structures. In SAS procedure “nlmixed”, any variance-covariance matrix must be manually coded.^12^ Though convergence issues are rare, our experience has taught us to save computational time and facilitate model convergence by taking the following actions: (i) Using the estimated model parameters from the MMRM model as the initial values for the proportional model. It is often sufficient to borrow only the estimated variance-covariance values as initial values, although both the estimated placebo mean decline and the variance-covariance matrix can also be borrowed. (ii) Similar to MMRM, if the model fails to converge using an unstructured variance-covariance matrix, simpler variance-covariance structures such as autoregressive order 1 can be employed to improve convergence. It is important to note from our experience that when a structured variance-covariance matrix is used, the sandwich estimator (“empirical” in SAS) should be employed to assist in controlling type I error. We recommend using the sandwich estimator even with an unstructured variance-covariance matrix when employing structured models such as proportional models or NCS models. In simulations, the estimation method must remain consistent for both power and type I error evaluation. If the sandwich estimator is used for one, it should also be used for the other.

Finally, these simulations highlight the impact of disease progression trajectory on model performance, which is expected. The placebo group consistently demonstrated decline across all post-baseline visits, regardless of the specific endpoint (e.g., CDR-SB, iADRS). However, certain endpoints, such as iADRS, ADAS-cog13, and logical memory delayed recall, exhibited a potential practice effect or trial effect (i.e., participants get better after enrolling in a trial), resulting in temporary improvement from baseline.^3, 8, 25^ In these cases, MMRM-based models utilizing all or multiple visits provide greater power without requiring additional model assumptions compared to the typical MMRM-Last-Visit model. In situations where the practice effect is not apparent, and the treatment exhibits disease-modifying effects, MMRM, pMMRM-based models using all or multiple visits, or pNCS are recommended due to their superior performance. Our simulation results indicate that once the last three post-baseline visits are included for efficacy inference, additional earlier post-baseline visits provide minimal gains in power. This is primarily due to a relatively lower signal-to-noise ratio in earlier visits, likely driven by cumulative treatment effects. This observation means that when a practice effect is evident in earlier post-baseline visits, proportional effect-based models can mitigate proportionality assumption violations by estimating the proportional effect using only the last three or even two visits. LME models, despite assuming strong linearity in disease progression and estimating fewer parameters, generally possess similar or lower power compared to MMRM/pMMRM-based models using all or multiple visits. Thus, LME models are not recommended in these cases. In some simulation scenarios, NCS demonstrates greater power compared to MMRM-Last-Visit. However, in other simulation scenarios, their power is similar. NCS is particularly suitable when dealing with messy data that is spread out relatively evenly across the trial duration, as documented in previous research.^18^ In the context of a typical randomized clinical trial, it is advisable to collect clinical or cognitive assessments based on a strict schedule. Such a schedule ensures data can reflect the disease progression from visit to visit (e.g., every 3 or 6 months per visit). These clinical trial data typically cluster around the scheduled visits. Consequently, it is reasonable to argue against analyzing data from a typical clinical trial using NCS, especially when disease progression trajectories are generally linear. In fact, the treatment effects estimated using MMRM in the TRAILBLAZER-ALZ 2 trial appeared consistently larger in magnitude compared to those estimated using NCS for various clinical endpoints in both the low/medium tau and the combined populations.^3^ When utilizing NCS as the primary analysis model, we highly recommend assessing the treatment efficacy by comparing the mean change from baseline, rather than relying on comparing the mean at the last visit. Our simulations have demonstrated that evaluating the mean change from baseline provides greater statistical power compared to mean at the last visit. Additionally, this approach is not impacted by any constraint placed on the baseline means.

In summary, the concept of proportional treatment effect has been widely established and extensively utilized for survival endpoints (e.g., Cox proportional hazard model^29^) and categorical endpoints (e.g., proportional odds ratio model^30^). These proportional models have significantly contributed to the advancement of treatment development across various medical disciplines. The use of similar proportional models such as pMMRM or pNCS to analyze continuous endpoints may have similar potential. Furthermore, the use of a proportional treatment effect extends its benefits beyond continuous endpoints. It provides the flexibility to simultaneously model multiple endpoints, including survival, categorical, or continuous, enhancing the power to detect an effective treatment effect across various domains.^12^ To empower researchers in implementing and comprehending these models, we have provided exemplary SAS codes that facilitate their implementation and understanding. By adopting models that exploit proportional treatment effects and/or incorporate all or multiple post-baseline visits, researchers can extract greater power from their clinical trial data, leading to increased efficiency in drug development. The application of these models is not limited to clinical trials for Alzheimer’s disease but can also be extended to other neurodegenerative diseases, such as Parkinson’s disease, frontotemporal lobe dementia, and amyotrophic lateral sclerosis.

## Data Availability

All data produced in the present work are contained in the manuscript

## Supplemental Materials

**Supplemental Table 1:** Proportional treatment effect (i.e., % reduction) at each visit of the TRAILBLAZER-ALZ 2 Donanemab Phase 3 Trial (low/medium tau population) in CDR-SB.

| Weeks | Estimated % Reduction | SE | 95% CI |  |
| --- | --- | --- | --- | --- |
|  |  |  | Lower | Upper |
| **12** | 36.4% | 24.1% | -10.9% | 83.7% |
| **24** | 48.1% | 11.8% | 25.0% | 71.3% |
| **36** | 51.3% | 8.4% | 34.8% | 67.9% |
| **52** | 43.9% | 7.0% | 30.1% | 57.8% |
| **64** | 38.3% | 6.7% | 25.2% | 51.5% |
| **76** | 36.2% | 5.9% | 24.6% | 47.8% |
| **% difference between Weeks 36 and 12** | 14.9% | 20.0% | -24.3% | 54.2% |
| **% difference between Weeks 36 and 64** | 13.0% | 7.1% | -1.0% | 27.0% |
| **% difference between Weeks 36 and 76** | 15.2% | 8.0% | -0.5% | 30.8% |
| These results were generated using simulated semi-real trial data, replicating the observed disease progression trajectory in the low-to-medium tau population of the TRAILBLAZER-ALZ 2 donanemab trial<sup>3</sup> |  |  |  |  |

**Supplemental Table 2:** Variance-covariance matrix and mean change from baseline (CFB) of CDR-SB extracted from Clarity-AD trial.

| Parameter | Months | 3 | 6 | 9 | 12 | 15 | 18 |
| --- | --- | --- | --- | --- | --- | --- | --- |
| Variance | 3 | 0.72 | 0.83 | 0.82 | 0.76 | 0.68 | 0.60 |
| Variance | 6 | 0.83 | 1.50 | 1.48 | 1.37 | 1.23 | 1.08 |
| Variance | 9 | 0.82 | 1.48 | 2.28 | 2.12 | 1.90 | 1.66 |
| Variance | 12 | 0.76 | 1.37 | 2.12 | 3.06 | 2.75 | 2.41 |
| Variance | 15 | 0.68 | 1.23 | 1.90 | 2.75 | 3.85 | 3.38 |
| Variance | 18 | 0.60 | 1.08 | 1.66 | 2.41 | 3.38 | 4.63 |
| Mean CFB | Placebo | 0.36 | 0.61 | 0.80 | 1.16 | 1.41 | 1.66 |
|  | Lecanemab | 0.27 | 0.43 | 0.58 | 0.81 | 1.02 | 1.21 |

**Supplemental Table 3:** Variance-covariance matrix and mean change from baseline (CFB) of CDR-SB extracted from the low-to-medium tau population of the TRAILBLAZER-ALZ 2 Trial.

| Parameter | Weeks | 12 | 24 | 36 | 52 | 64 | 76 |
| --- | --- | --- | --- | --- | --- | --- | --- |
| Variance | 12 | 1.76 | 1.59 | 1.40 | 1.25 | 1.07 | 0.90 |
| Variance | 24 | 1.59 | 2.24 | 1.98 | 1.76 | 1.50 | 1.28 |
| Variance | 36 | 1.40 | 1.98 | 2.72 | 2.42 | 2.07 | 1.76 |
| Variance | 52 | 1.25 | 1.76 | 2.42 | 3.37 | 2.88 | 2.45 |
| Variance | 64 | 1.07 | 1.50 | 2.07 | 2.88 | 3.85 | 3.27 |
| Variance | 76 | 0.90 | 1.28 | 1.76 | 2.45 | 3.27 | 4.34 |
| Mean CFB | Placebo | 0.28 | 0.64 | 0.95 | 1.35 | 1.53 | 1.88 |
|  | Donanemab | 0.18 | 0.33 | 0.46 | 0.75 | 0.94 | 1.20 |

**Supplemental Table 4:** Variance-covariance matrix and mean change from baseline (CFB) of iADRS extracted from the low-to-medium tau population of the TRAILBLAZER-ALZ 2 Trial.

| Parameter | Weeks | 12 | 24 | 36 | 52 | 64 | 76 |
| --- | --- | --- | --- | --- | --- | --- | --- |
| Variance | 12 | 77.07 | 63.73 | 52.59 | 43.73 | 35.94 | 29.50 |
| Variance | 24 | 63.73 | 82.34 | 67.95 | 56.50 | 46.44 | 38.12 |
| Variance | 36 | 52.59 | 67.95 | 87.61 | 72.85 | 59.88 | 49.15 |
| Variance | 52 | 43.73 | 56.50 | 72.85 | 94.65 | 77.80 | 63.86 |
| Variance | 64 | 35.94 | 46.44 | 59.88 | 77.80 | 99.92 | 82.02 |
| Variance | 76 | 29.50 | 38.12 | 49.15 | 63.86 | 82.02 | 105.19 |
| Mean CFB | Placebo | -0.91 | -1.79 | -3.29 | -5.26 | -7.03 | -9.61 |
|  | Donanemab | 0.48 | 0.30 | -0.94 | -2.74 | -3.50 | -5.81 |

